# Comparison of 11 High Frequency Oscillation (HFO) Detectors across Scalp and Intracranial EEG to Evaluate Clinical Utility

**DOI:** 10.64898/2026.05.05.26352459

**Authors:** Margarita Maltseva, Daniel Lachner-Piza, Pierre LeVan, Minette Krisel Manalo, Walter Hader, Julia Jacobs

## Abstract

**Introduction:** To leverage high-frequency oscillations (HFOs) as a biomarker with significant potential, this study compared a large set of detectors on a unified dataset, aiming to evaluate their clinical applicability under realistic conditions.

**Methods:** Eleven automatic detectors were applied to a retrospective dataset of intracranial and scalp EEGs from 27 consecutive pediatric patients. Inter-detector agreement was assessed using Spearman’s Rho, and the area under the curve (AUC) for seizure onset zone (SOZ) prediction served as a consistent reference standard to enable reliable comparisons across recording modalities. Analyses were conducted separately for HFO and Spike-HFO detections.

**Results:** The average age of our cohort was 12.4 years (SD 4.0; range 5–18). AUC values in scalp EEG ranged from 0.61 to 0.67 for HFOs and from 0.53 to 0.63 for Spike-HFO. AUC values in intracranial EEG ranged from 0.48 to 0.66 for HFOs and 0.54 to 0.69 in Spike-HFO. Although only three of the 11 detectors were specifically developed or adapted for scalp EEG, the detectors generally achieved higher AUC values and stronger agreement in scalp EEG

**Conclusions:** We present the first study comparing intracranial and scalp detectors by testing them beyond the modalities for which they were originally designed. Although the clinical utility of detections was comparable across EEG modalities, it remained lower than reported in original studies assessing the diagnostic value of HFOs. Caution is warranted when applying a publicly available detector to a new dataset, and detector robustness remains a critical issue.

**Key points:**

- A comprehensive head-to-head comparison of 11 detectors demonstrated significant variability in detector agreement and clinical utility
- Clinical utility was not necessarily linked to the EEG recording type the detector was originally designed for
- Despite widely accepted use of automatic detections, detector robustness remains a critical issue

## 1.

High frequency oscillations (HFOs) have emerged as a valuable diagnostic tool, complementing established EEG markers such as spikes in intracranial and scalp EEG.^1–3^ A common definition, applied to this study, states that HFOs are spontaneous EEG patterns within the 80–500 Hz range, consisting of at least four oscillations clearly distinguishable from the background and characterized by a typical duration of 30–100 ms and an inter-event interval of at least 25 ms.^4^

Despite more than two decades of HFO research, accurate event identification remains a challenge. Visual marking by trained neurophysiologists is time-consuming and subject to high interrater variability, making automatic detection essential for both systematic research and clinical application.^5^ However, particularly in scalp EEG, the precision and reliability of automatic HFO detectors remain underexplored and consequently compromises the clinical utility of the biomarker.^6^ Moreover, existing detectors vary widely in their methodology.^6^ Additionally, HFO definitions are not yet standardized, and visual marking in scalp EEG is further complicated by artifacts, localization difficulties, and low signal-to-noise ratios.^7, 8^ Addressing these challenges could enable broader application, supporting presurgical evaluation, disease monitoring, and seizure prediction.

Most detection algorithms are developed and tested on datasets collected at a single site, leading to good intra-dataset performance but poor generalizability across different datasets. Preprocessing steps often optimize the signal-to-noise ratio, but this limits detectors’ ability to manage novel artifacts and noise, which can yield inconsistent results. Modifying algorithms for specific datasets by adjusting filters and thresholds requires extensive expertise in signal processing, making it impractical for routine clinical use.^4^ The rate of HFO in a channel seems a reliable marker of underlying epileptogenic tissue, and it has been the most commonly used metric.^8^ Detection algorithms are often validated by visual correction; however the clinical implications of the automatic detections remain uncertain. In any case, the ultimate validation will be given by the usefulness of the detections.^8^

To address this gap, this study aims to compare the clinical utility of a large set of HFO detectors on a single dataset, using the default parameters specified in their original publications. To evaluate the diagnostic value across different recording types – scalp and intracranial EEG – the seizure onset zone (SOZ) localization was used as a consistent gold standard to ensure reliable comparisons.

## 2. Methods

### 2.1 Patient selection

Consecutive patients recorded between 2019 and 2024 at the Alberta Children’s Hospital were included in this analysis. Data collection was performed retrospectively by retrieving files from the clinical EEG database. Due to the patient collection site, eligible patients ranged in age from 0 to 18 years. Inclusion criteria were a clinical diagnosis of epilepsy, an existing intracranial monitoring with a sampling rate > 1 kHz, and a clearly identifiable seizure onset zone (SOZ) in the EEG. Patients were excluded if the presence of artifacts rendered the analysis infeasible or if the SOZ was defined as diffuse by a multidisciplinary consensus (consisting of multiple epileptologists and neurosurgeons, as per institutional guidelines). The study was approved by the local research ethics board (REB20-0938).

### 2.2 EEG segment selection

For scalp EEG, a long-term video-EEG recording with 10–20 electrode configuration, conducted prior to the intracranial monitoring, was selected for each patient. Intracranial monitoring was performed according to best clinical practices, with the implantation sites previously decided by a multidisciplinary team. For each patient and recording type, 30-minutes of unfiltered EEG were selected for automatic analysis. Selection criteria favoured sleep stage II, minimal artifacts, and the absence of electroclinical seizures. The selected EEG segments were at least one hour apart from electro-clinical seizures. The reviewers (MM, MKM) excluded any EEG sections with obviously visible artifacts when clipping the files. When possible, the first night of recording was avoided for intracranial EEGs to minimize the interference from the implantation procedure.

### 2.3 Automatic Detectors

To detect spikes, we employed the Delphos-Spikes-detector.^9^ A total of 11 different HFO detectors were then evaluated in this comparison. Each detector was run using the default parameters as specified in the publications mentioned in Table 1.

**Table 1.** Overview of implemented detectors with each defining publication and detector characteristics.

| **Intracranial HFO detectors** |  |  |
| --- | --- | --- |
| Defining publication | Detector characteristics | Referred to as |
| Zelmann et al. 2012<sup>4</sup> and implemented in Navarrete et al. 2016<sup>10</sup> | Wavelet entropy baseline detector | MNI-RZ |
| Crépon et al. 2010<sup>11</sup> and implemented in Navarrete et al. 2016<sup>10</sup> | Uses the Hilbert Transform to compute the envelope of the filtered signal | HIL |
| Gardner et al. 2007<sup>12</sup> and implemented in Navarrete et al. 2016<sup>10</sup> | Short Line Length detector | SLL |
| Staba et al. 2002<sup>13</sup> and implemented in Navarrete et al. 2016<sup>10</sup> | Short Time Energy detector | STE |
| Roehri et al. 2016<sup>9</sup> and implemented in Roehri et al 2017<sup>14</sup> | Time-frequency based detector with prewhitening | Delphos-cmd |
| Staba et. al 2002<sup>13</sup> and implemented in Gliske et al. 2015<sup>15</sup> | Thresholding Root mean square detector | Michigan |
| Ellenrieder et al. 2012<sup>16</sup> and implemented in von Ellenrieder et al. 2016<sup>17</sup> | Thresholding Root mean square detector | MNI-NVE |
| Lachner-Piza et al. 2020<sup>18</sup> and implemented in Lachner-Piza et al. 2020<sup>18</sup> | Based on rbf kernel support-vector-machines | MOSSDET |
| **HFO Detectors originally developed or adjusted for scalp EEG** |  |  |
| Defining publication | Detector characteristics | Referred to as |
| Chu et al. 2017<sup>19</sup> and implemented in the author's repository (https://github.com/Mark-Kramer/Spike-Ripple-Detector-Method/tree/master) | Computes the amplitude envelope to identify spike-ripple events | Harvard |
| Charupanit and Lopour 2017<sup>20</sup> | Amplitude threshold-based detector with an iterative approach | Cali-Irvine |
| Fedele et al. 2016<sup>21</sup> and implemented in Fedele et al. 2017<sup>22</sup> | Time-frequency based detector | Zurich |

### 2.4 Data analysis

Since the HFO-generating area is typically defined by the EEG channels with the highest HFO occurrence rates, we selected Spearman’s rank correlation as the most clinically relevant metric to assess agreement between detectors. Agreement was calculated by comparing EEG channels according to their HFO rate per minute and calculating the resulting Spearman’s rank correlation coefficient (ρ) between each pair of detectors. Spearman’s Rho values were computed for both HFOs alone and for the subset of HFOs that were temporally and spatially coincident with interictal epileptic spikes (Spike-HFOs). Spikes were detected using the Delphos-Spikes detector.^9^ Correlation values were interpreted as follows: ≥ 0.7 (very strong relationship), 0.4-0.69 (strong relationship), 0.3-0.39 (moderate relationship), and 0.2-0.29 (weak relationship).^23^

The utility of HFO and Spike-HFO detections as epilepsy biomarkers was compared between detectors based on their ability to localize the SOZ, defined by a multidisciplinary consensus of multiple epileptologists, neuropsychologists, and neurosurgeons. Study eligibility was verified by an expert epileptologist (JJ).

The ability of HFO and Spike-HFO to localize the SOZ was assessed by generating a receiver-operating-characteristic (ROC) curve, which measures the sensitivity and the false-positive-rate (1 - specificity) obtained when iterating through a range of thresholds on the average occurrence rates of HFO and Spike-HFO in each channel across the entire EEG segment. The area under the ROC curve (AUC) was used as the summarizing metric of each detector’s success in localizing the SOZ. The 95% confidence interval was computed using bootstrapping with 10000 samples with replacement and the mean as the aggregating metric. All analyses were performed separately for intracranial and scalp EEG recordings.

## 3. Results

### 3.1 Clinical and demographic characteristics

In total, 27 consecutive patients were included in this study, who underwent separate intracranial and scalp EEG studies. The average age at the time of intracranial EEG recording was 12.4 years (SD 4.0; range 5-18) and 46.4% of our study population was female. All patients had a diagnosis of focal epilepsy with the following underlying etiologies: 29.6% focal cortical dysplasia (FCD) (n=8), 22.2% unknown etiology (n=6), 14.8% hippocampal sclerosis (n=4), 11.1% tuberous sclerosis (n=3), 11.1% subpial gliosis (n=3), 3.7% remote ischemic event (n=1), 3.7% polymicrogyria (n=1), 3.7% dysembryoplastic neuroepithelial tumor and hippocampal sclerosis (n=1).

### 3.2 Agreement between detectors in scalp EEG

For scalp EEG, the detectors showed strong to very strong with ρ values up to 0.89 (Figure 2A). The Zurich and Harvard detectors showed comparatively lower agreement, with lower ρ values. Agreement in Spike-HFO detection was generally lower, with ρ values ranging from 0.15 to 0.65. The Zurich and Harvard detectors showed again a lower agreement between Spike-HFOs compared to other detectors.

**Figure 1.**
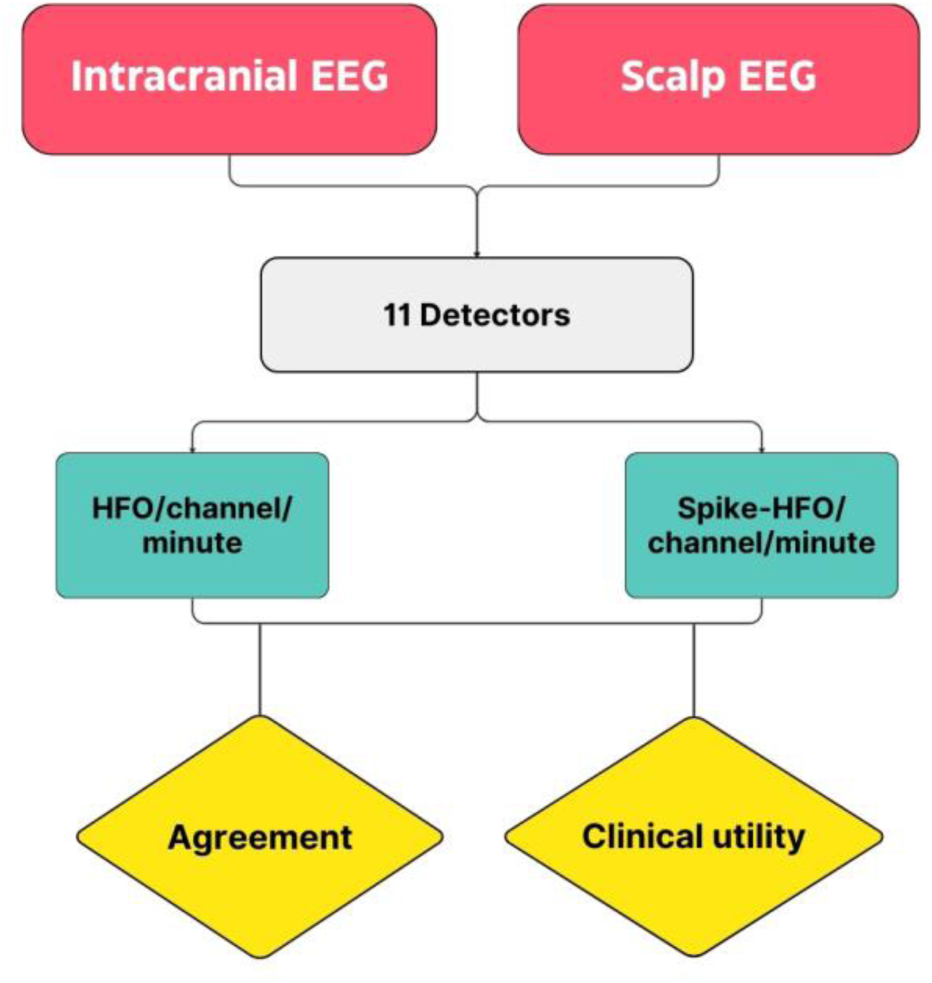
Study design. Two different EEG recording types were analyzed by 11 detectors for HFO and Spike-HFO rates. Event rates were then used to compare detector agreement and clinical utility.

**Figure 2.**
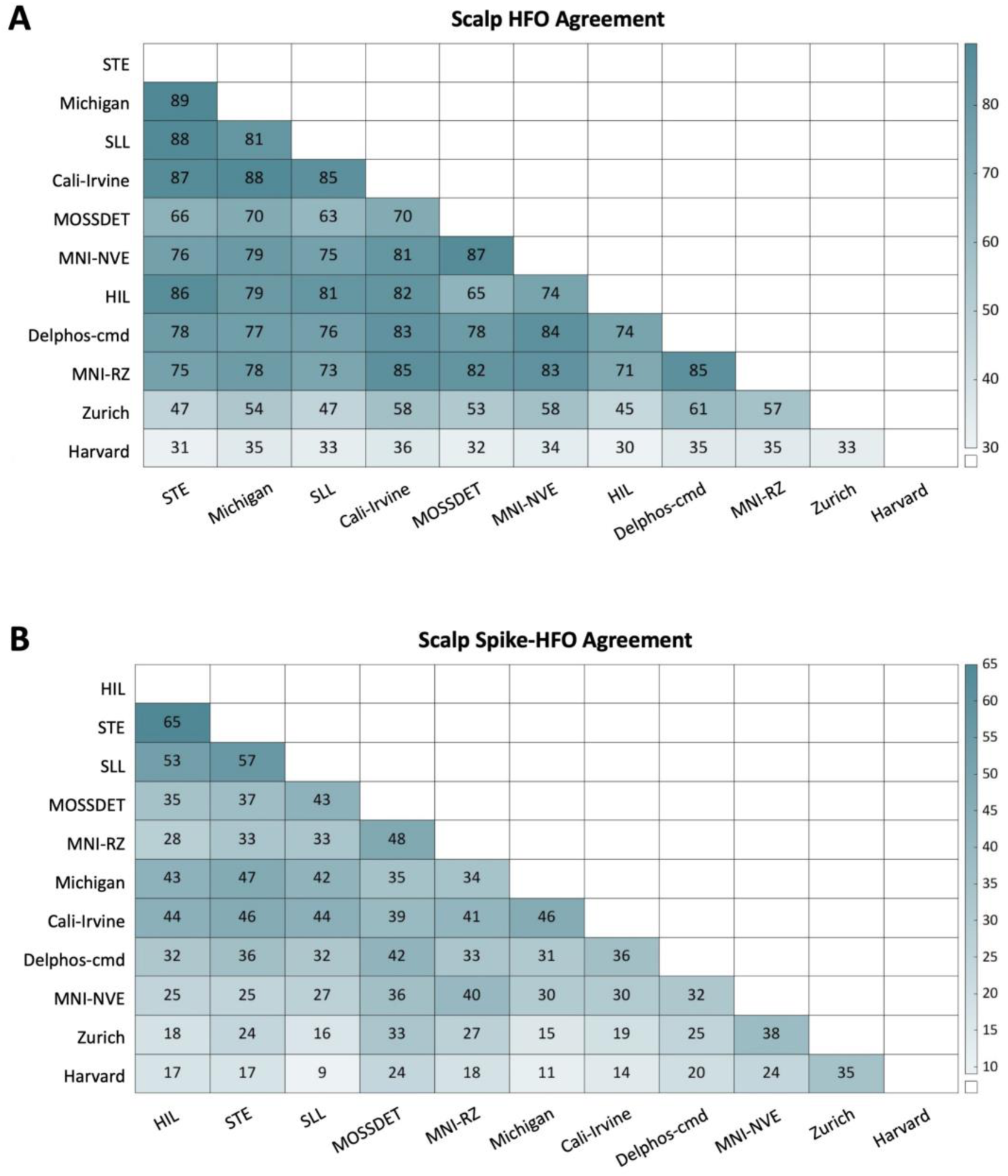
Spearman’s Rho Correlation Coefficient assessing the agreement between detectors in scalp EEG, sorted in descending order of average agreement. The scale shows ρ values multiplied by one hundred. Darker colour indicates stronger agreement. **A**. Agreement on HFO detections. **B**. Agreement on Spike-HFO detections.

### 3.3 Agreement between detectors in intracranial EEG

In intracranial EEG, ρ values ranged from −0.18 to 0.87, indicating a broader variance in agreement (Figure 3). The MNI-RZ detector showed lower agreement, while the Zurich and MOSSDET detectors achieved the highest agreements (0.87), closely followed by Delphos-cmd and MNI-NVE detectors (0.86). Agreement for Spike-HFOs in intracranial EEG (Figure 3B) was generally higher than for HFOs (Figure 3A), with ρ values ranging from 0.40 to 0.84, except for the Michigan and MNI-RZ detectors.

**Figure 3.**
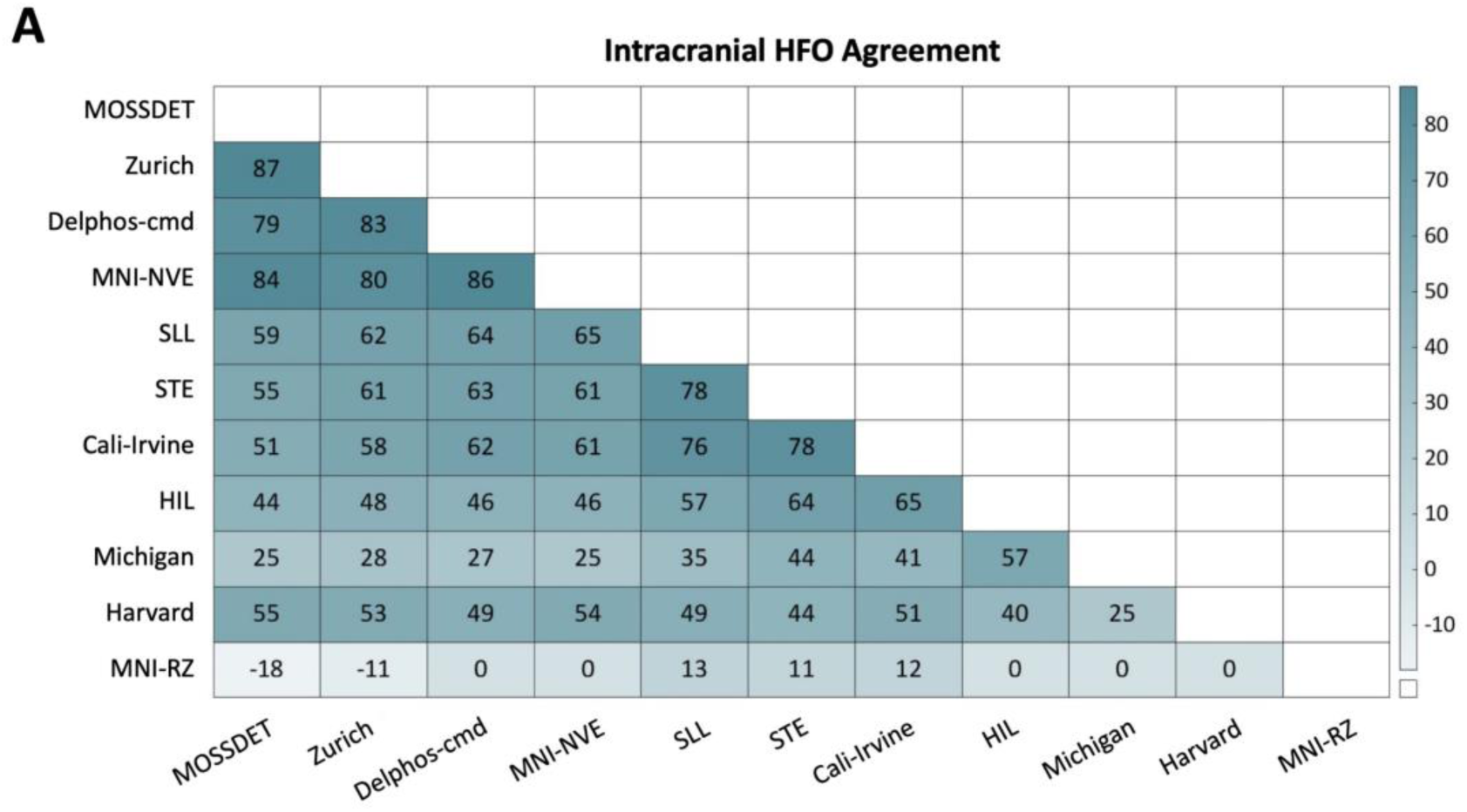
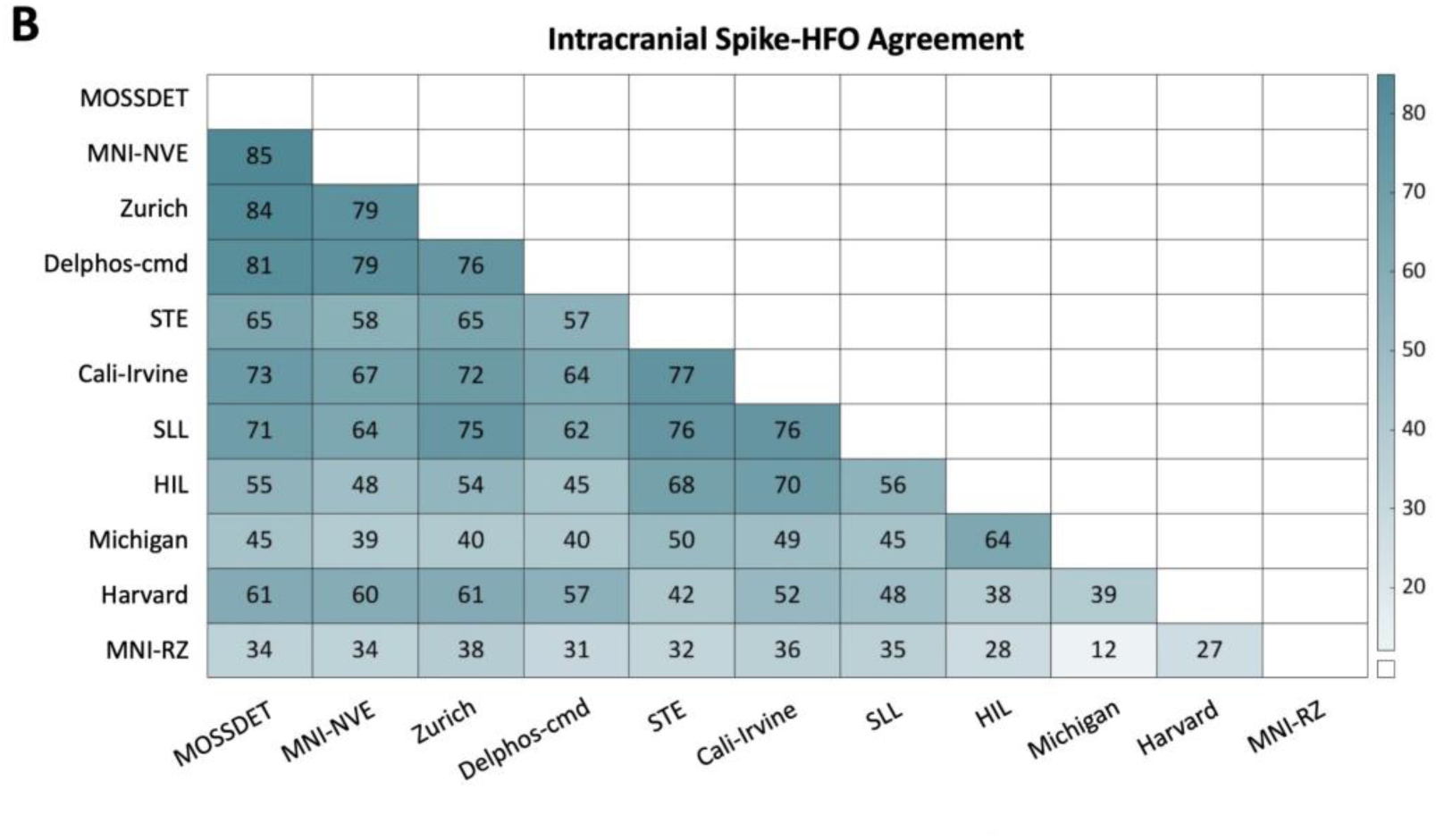
Spearman’s Rho Correlation Coefficient assessing the agreement between detectors in intracranial EEG, sorted in descending order of average agreement. The scale shows ρ values multiplied by one hundred. Darker colour indicates stronger agreement. **A**. Agreement on HFO detections. **B**. Agreement on Spike-HFO detections.

### 3.4 Predictability of the seizure onset zone (SOZ)

A random classifier was computed (dotted line in the ROC diagrams), representing a performance baseline. ROC curves were additionally calculated for Delphos-Spikes, to assess whether HFOs or Spike-HFOs provided an enhanced biomarker value.

For scalp EEG, the area under the curve (AUC) for HFOs ranged from 0.61 to 0.67, with the highest AUC achieved by the MNI-NVE detector. The AUC for Spike-HFOs in scalp EEG ranged from 0.53 to 0.63, with the Delphos-cmd detector showing the highest AUC value. With 0.69, the overall highest AUC in scalp EEG was obtained with the Delphos-Spikes detector (Figure 4)

**Figure 4.**
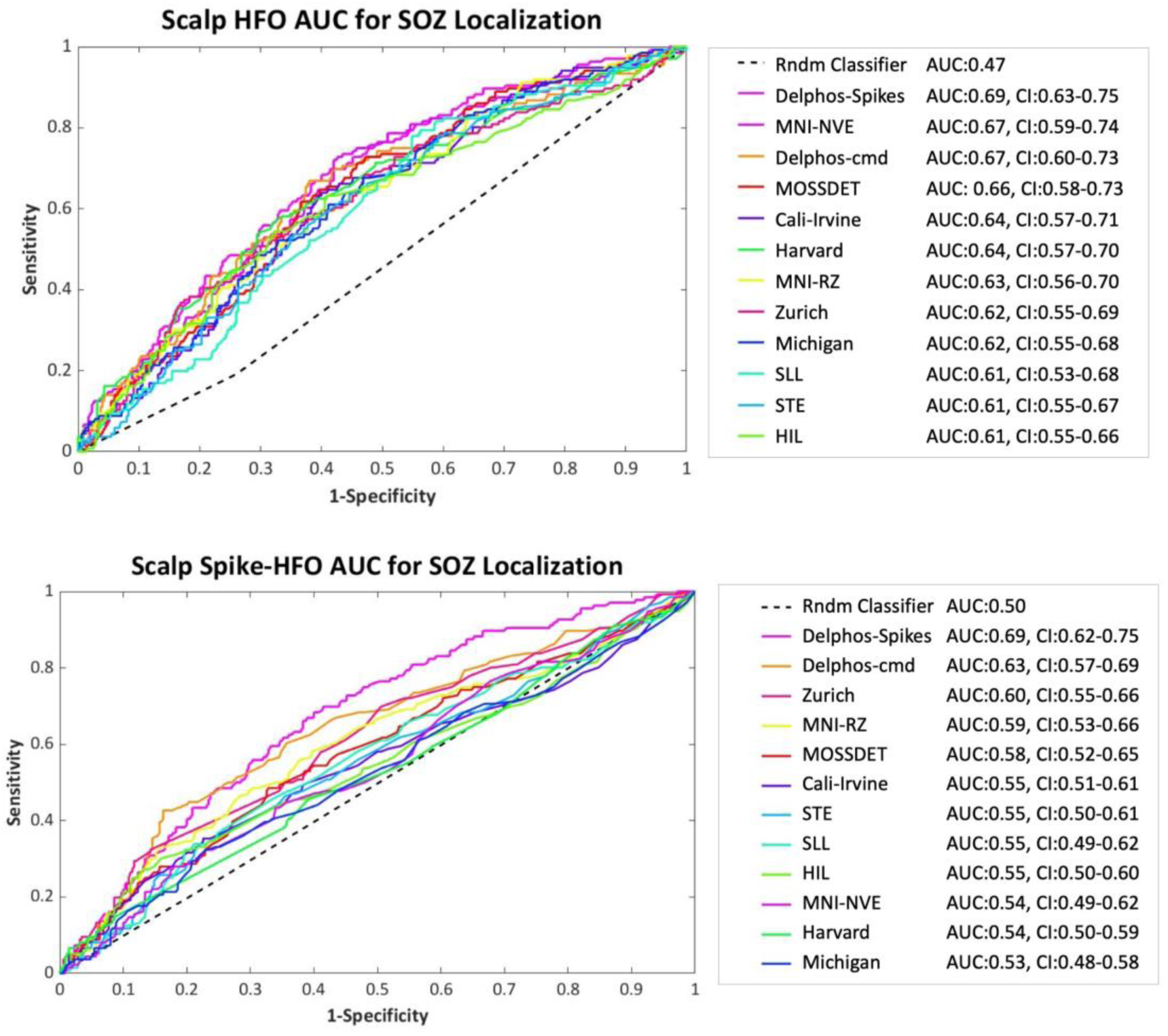
Clinical utility of scalp EEG detections. Clinical utility of the detections was assessed based on the seizure onset zone localization (SOZ). The area under the receiver operating characteristic curve (AUC) was calculated for HFO, Spike-HFO and spike detections (Delphos-Spikes). The dotted line represents AUC values of a random classifier (Rndm Classifier).

For intracranial EEG, the AUC for HFOs ranged from 0.48 to 0.66, with the Cali-Irvine detector achieving the highest value. The AUC for Spike-HFOs in intracranial EEG ranged from 0.54 to 0.69, with the highest value observed using the MOSSDET detector. With 0.69, the overall highest AUC in intracranial EEG was obtained with the Delphos-Spikes detector (Figure 5).

**Figure 5.**
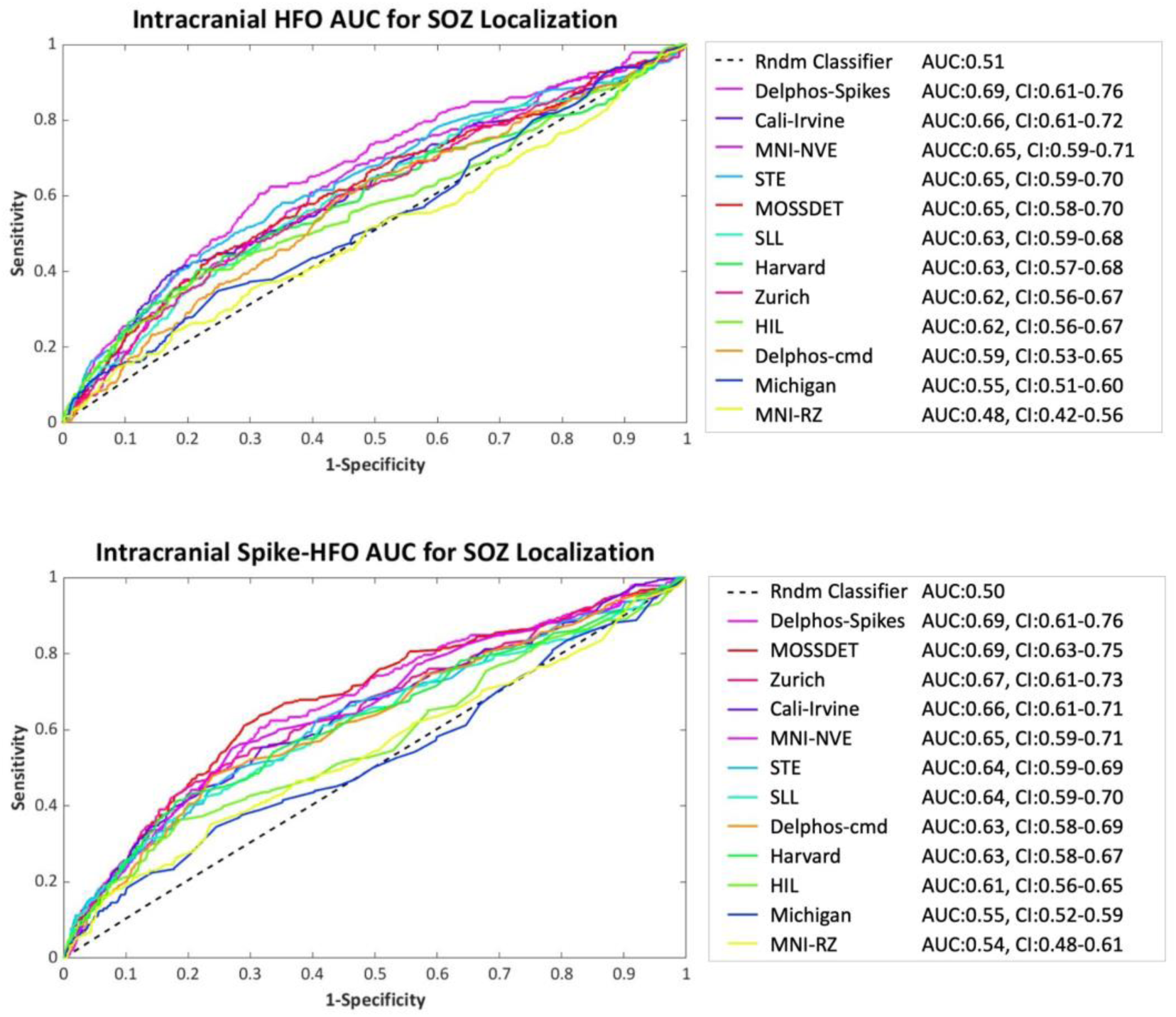
Clinical utility of intracranial EEG detections. Clinical utility of the detections was assessed based on the seizure onset zone localization (SOZ). The area under the receiver operating characteristic curve (AUC) was calculated for HFO, Spike-HFO and spike detections (Delphos-Spikes). The dotted line represents AUC values of a random classifier (Rndm Classifier).

## 4. Discussion

We present a comprehensive head-to-head comparison of 11 automatic HFO detectors on a unified dataset, and the first study comparing intracranial and scalp detectors by testing them beyond the modalities for which they were originally designed.

Our results demonstrate considerable variability in the clinical utility of the detections and inter-detector agreement, mirroring the similar challenges seen in visual HFO marking.^16, 24^ The variability in clinical utility was not necessarily linked to EEG recording type (scalp or intracranial). Although some detectors displayed high agreement, none achieved an AUC value greater than 0.69 in predicting the seizure onset zone (SOZ).

While all detectors performed better than a random classifier in SOZ prediction, caution should be applied when implementing a publicly available detector on a new data set.

### 4.1 Clinical utility does not necessarily correlate with EEG recording type

Interestingly, the clinical utility of the detections did not necessarily correlate with the EEG recording type for which each respective detector was originally designed. The most significant concern for intracranially developed detectors used on scalp EEG without adaptation would be contamination from artifacts generated by craniofacial muscle activity. Although only three of the 11 detectors were specifically developed or adapted for scalp EEG,^19–22, 25^ the majority of the detectors achieved higher AUC values and stronger agreement in scalp EEG. This underscores the potential of HFO as a non-invasive biomarker, despite the possibility of artifacts from high-frequency signals that could heavily obscure the ripple band.^26^

Given the lack of a formal definition of HFOs, comparing detectors within a single dataset is important to assess them in a realistic setting. Previous literature suggests validating HFO detectors based on visual marking or human review as a gold standard.^4^ However, from a clinical perspective, it is more relevant to validate detections based on their utility in addressing a diagnostic question, such as defining the SOZ.^8^ Successful application of the biomarker could possibly be achieved when similar conclusions in terms of epileptogenic regions or cognitive function location are obtained by the automatic methods as compared to the human experts.^8^ Even more striking would be if conclusions from single HFO studies, promising high diagnostic value, can be reproduced in a realistic setting.

A systematic review and meta-analysis, conducted by Wang et al., on the prognostic value of intracranial HFOs defining the SOZ based on complete surgical resection, showed pooled AUC values between 0.76-0.81, which is substantially higher than any of the values calculated in our sample.^27^ Our first hypothesis is that HFOs could be more strongly correlated with resected tissue than the SOZ only. Another hypothesis could be that the detections in this study were affected by false positives. Since surgical candidacy was not a selection criterion for this study, it is not possible to test the first hypothesis, which is a limitation of this manuscript. The potential for false positives will be discussed in more detail below.

### 4.2 Factors influencing HFO detections differently between EEG recording types

When comparing EEG modalities, an overall higher agreement among HFO only was observed in scalp EEG, whereas intracranial EEG showed a stronger agreement between Spike-HFOs. The inter-detector highlights the influence of similarities in the detectors’ underlying approaches. Compared to intracranial detectors, two scalp detectors (Zurich and Harvard), demonstrated lower agreement on scalp EEG, but moderate agreement when compared only between each other. This could potentially be explained by a lower susceptibility to high frequency noise compared to the rest of the detectors. The Zurich detector identifies ripple events using a time-frequency baseline and an amplitude envelope threshold. Similarly, the Harvard detector uses an envelope threshold to detect ripple events; however, it aims to identify ripple events that co-occur with interictal spikes. The third detector that is also designed for scalp EEG (Cali-Irvine) has a similar approach to identify potential ripple events; it calculates an optimal amplitude threshold based on the amplitude of each oscillation in an iterative process. Despite the similarity in approach, this detector showed an overall higher agreement with detectors designed for intracranial EEG recordings than with the two scalp detectors.

One key aspect that might influence detection results differently between scalp and intracranial EEG is that scalp HFOs are more frequently visible in the ripple range (80-250Hz). Therefore, scalp detectors are likely designed to identify HFOs in this frequency range and reject events in the surrounding frequencies. In contrast, intracranial detectors might be more sensitive to identify distinct oscillations in the frequency range above 250Hz, but also more prone to detect high frequency noise as HFOs.^28^

The only way to conclude on whether detectors agree on false positives would be through visual validation of the detections, which is corroborated by recent data. A study evaluating ripple band markings by the Cali-Irvine detector found only 43.2% of detections to be true HFOs, with 55.1% classified as background activity and 1.7% as artifacts.^29^ They also reported that the percentage of true HFOs varied significantly between patients. Similarly, von Ellenrieder et al. reported high rates of false positives per minute at 95% detection sensitivity in scalp EEG.^16^ A study by Charupanit et al. tested a detector originally designed for intracranial EEG, but adapted for the detection of scalp HFOs, and confirmed that the percentage of events classified as artifacts or background varied randomly between datasets. To resolve this issue, they implemented four post-processing rejection steps and human validation.^30^

Overall, a lower signal-to-noise ratio as well as a small amplitude of scalp HFOs might decrease the sensitivity and specificity in automatic detection, demanding a semi-automatic approach.^31^ At the same time, future researchers have to be mindful that visual analysis is not perfect, as identifying events in a high frequency background is challenging for the human eye. Nonetheless, the potential of HFOs as a non-invasive biomarker is immense, and reliably adapting detectors to scalp EEG is a crucial step in exploring this potential.

### 4.3 Improving detector robustness remains crucial to enabling clinical application

Another potential explanation for the variability in detections lies in the characteristics of the dataset. Each algorithm was run with its published default parameters, but because our recordings come from pediatric patients with therapy-refractory focal epilepsy, they may share similarities with those used to develop and train some of the detectors. This could influence performance and therefore the clinical utility resluts of the detections. For example, the MOSSDET detector achieved the highest AUC with intracranial Spike-HFO detections. MOSSDET is the detector that we normally use in our lab, and thus likely most adapted to the dataset used in this study. In line with this idea, previous studies report significant performance improvements following threshold optimization tailored to specific datasets.^4, 30^ That said, preprocessing EEG data and adjusting detector parameters require expertise and considerable time effort, posing a significant burden to realistic clinical settings. Extensive HFO research of the past decades led to increased trust in the biomarker and detection methodologies. Consequently, we recommend that initial detector implementation should at least include some visual inspection before unsupervised use. We recognize that verifying every single event may not be feasible or valuable, due to inherent limitations of visual analysis. Incorporating artifact rejection tools – either as part of the detector or as a preprocessing step – could enhance applicability.

Particularly concerning is a growing body of literature showing that openly available detectors are being applied to new datasets without specifying any adaptation or validation results.^32, 33^ Even though those automatic detectors are previously validated by their developer, the performance results do not necessarily translate to the new data set. Hence, our findings emphasize that improving detector robustness to handle variability in EEG recordings remains a crucial issue that must be addressed to enable the clinical application.

### 4.4 Comparing clinical utility and resulting challenges

Previous research suggests that Spike-HFOs localize the SOZ with higher specificity than HFOs or spikes alone.^7, 16, 34–36^ Consistent with this, slightly higher AUC values were observed for Spike-HFO detections in intracranial EEG, suggesting greater pathological significance. Interestingly, spikes detected by the Delphos-Spike-detector still achieved higher AUC values than HFOs in intracranial EEG. Although overall similar to intracranial AUC values, the highest AUC in scalp EEG was also obtained with the Spikes only detector, once more questioning the clinical utility of the other detections.

It is important to discuss that even though a clear SOZ was a selection criterion for this study, an inherent limitation of scalp EEG is the spatial resolution and therefore the ability to precisely identify the SOZ. It is also worth considering whether the SOZ is the most accurate marker. Accumulating evidence suggests that epilepsy arises from a network disorder, and HFOs within this network may hold more diagnostic significance than those identified solely within the SOZ.^37^ With HFOs becoming more popular as a non-invasive biomarker their clinical utility expands towards measuring disease activity, treatment success and even epileptogenesis.^38–40^ Furthermore, physiological HFOs can complicate the evaluation of detections, especially, as their prevalence varies among patients.^41, 42^ Fast ripples are less likely to mimic physiological HFOs, but require higher sampling rates and have little applicability in scalp EEG.^43, 44^

Finally, this manuscript does not claim to provide a complete comparison of all available detection algorithms that have been detailed in the literature. We acknowledge that there might be more recent or higher-performing HFO detectors, which biases general conclusions about detector performance.^45–47^ Nonetheless, many of the presented algorithms are still in use.^48, 49^ In the previous literature, the rate of HFO in a channel seems to be a reliable marker of underlying epileptogenic tissue, and it has been the most commonly used metric.^8^ However, other measures – such as ranking of channel durations, amplitudes, peak frequency, temporal distributions, and entropy – might turn out to be more appropriate to characterize HFO activity.^8^ Newer methods, including advanced signal processing and deep learning, provide a promising opportunity to address the outlined issues and transfer HFO application into clinical routine.^50^

## 5.

This is the first study to provide a comprehensive comparison of 11 automatic HFO detectors while using them on a single dataset of intracranial and scalp EEG, highlighting the variability and challenges in automatic HFO detection. While detectors demonstrated promising performance, no single detector consistently excelled across all metrics or EEG types. Our findings emphasize the importance of applying caution when employing detectors to new datasets. Despite the widely accepted use of automatic HFO detectors, improving detector robustness remains a critical step toward clinical applicability.

## Data Availability

Since the EEG data files include sensitive patient information and therefore belong to the Alberta Childrens Hospital, uploading them to a public repository could compromise individual privacy. All other data produced in the present study are available upon reasonable request to the authors (submitted to ORCID 0000-0002-8598-4357).

## Contributions

MM has contributed to the conception of the work as well as the acquisition, analysis, and interpretation of data. MM has also drafted the work and revised it critically for important intellectual content.

DLP has contributed to the conception of the work as well as the acquisition, analysis, and interpretation of data. DLP has also revised the work critically for important intellectual content and contributed to the final approval of the version to be published.

PL has contributed to the interpretation of data, revised the work critically for important intellectual content, and contributed to the final approval of the version to be published.

MKM has contributed to the acquisition of data and contributed to the final approval of the version to be published.

WH has contributed to the acquisition of data and to the final approval of the version to be published.

JJ has contributed to the conception of the work as well as the acquisition, analysis, and interpretation of data. JJ has also revised the work critically for important intellectual content and contributed to the final approval of the version to be published.

